# Fractal kinetics of COVID-19 pandemic (with update 3/1/20)

**DOI:** 10.1101/2020.02.16.20023820

**Authors:** Anna L. Ziff, Robert M. Ziff

## Abstract

We give an update to the original paper posted on 2/17/20 – now (as of 3/1/20) the China deaths are rapidly decreasing, and we find an exponential decline to the power law similar to the that predicted by the network model of Vazquez [2006]. At the same time, we see non-China deaths increasing rapidly, and similar to the early behavior of the China statistics. Thus, we see three stages of the spread of the disease in terms of number of deaths: exponential growth, power-law behavior, and then exponential decline in the daily rate.

(Original abstract) The novel coronavirus (COVID-19) continues to grow rapidly in China and is spreading in other parts of the world. The classic epidemiological approach in studying this growth is to quantify a reproduction number and infection time, and this is the approach followed by many studies on the epidemiology of this disease. However, this assumption leads to exponential growth, and while the growth rate is high, it is not following exponential behavior. One approach that is being used is to simply keep adjusting the reproduction number to match the dynamics. Other approaches use rate equations such as the SEIR and logistical models. Here we show that the current growth closely follows power-law kinetics, indicative of an underlying fractal or small-world network of connections between susceptible and infected individuals. Positive deviations from this growth law might indicate either a failure of the current containment efforts while negative deviations might indicate the beginnings of the end of the pandemic. We cannot predict the ultimate extent of the pandemic but can get an estimate of the growth of the disease.

## 1 Update 3/1/20

Since the original posting of this paper on February 17 (which we are leaving intact below for comparison), the epidemic has fortunately slowed down considerably in China, and the current number of deaths there is 2,873 (3/1/20). At the same time, the number of deaths outside of China (equal to 104) is growing very rapidly, almost exponentially. Therefore we now separate out the China cases from the non-China cases.

In Fig. 1 is a log-log plot of the deaths in China, showing a fit of the power-law behavior we saw before, fit for the days 1/28 to 2/16. It can be seen that the power-law we saw before continued for a few more days until about 2/20, at which point the mortality began to decrease markedly.

**Figure 1:**
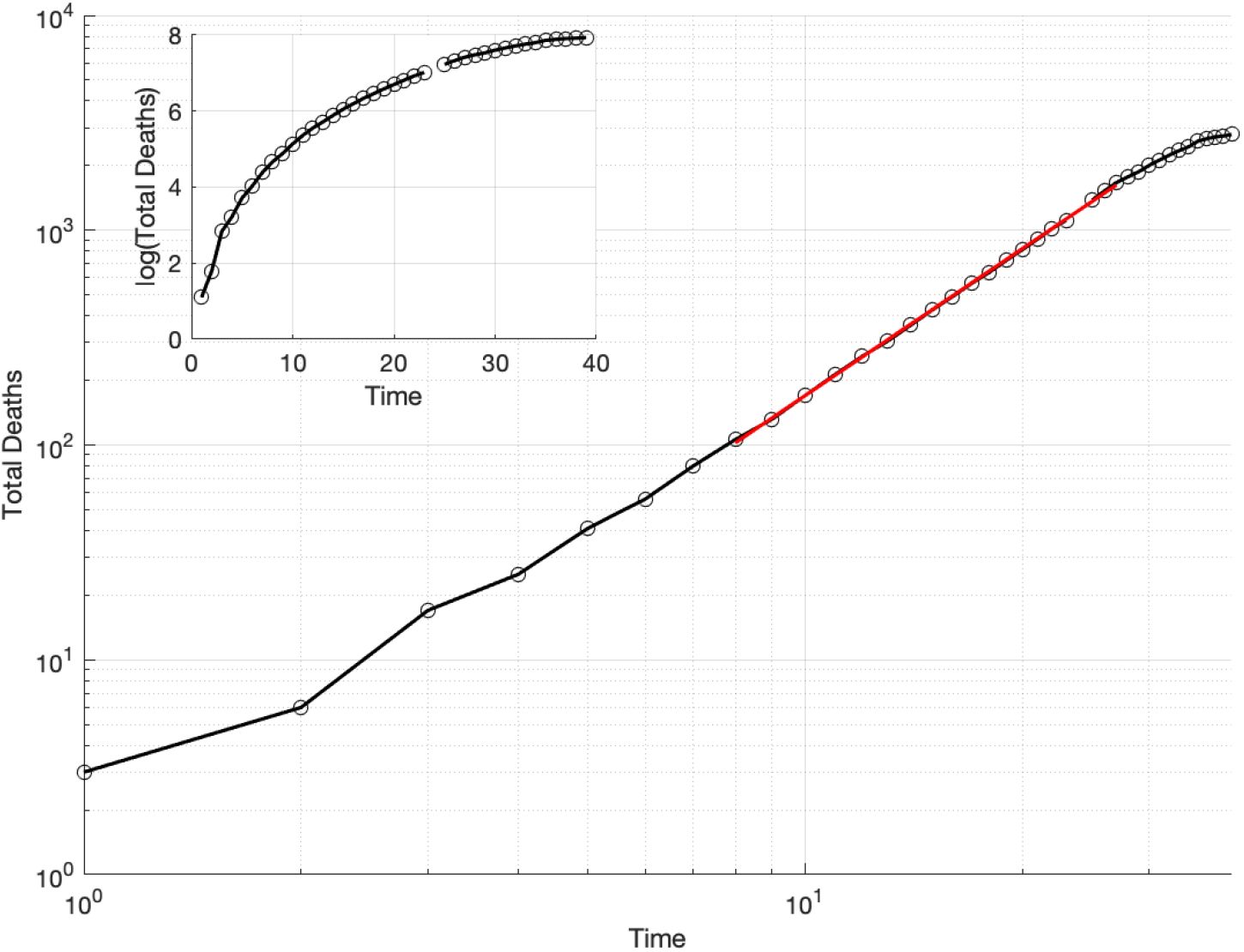
Deaths in China vs. Time (days since 1/21) on a log-log plot), and (inset) on a log-linear plot. The line is fit through the points filled in red, from day 8 (1/28) to day 27 (2/16). Data from World Health Organization [2020].

In Fig. 2 we show the number of deaths in China each day, now plotted on a log-linear plot, and we see a sharp cutoff during the last week or so.

**Figure 2:**
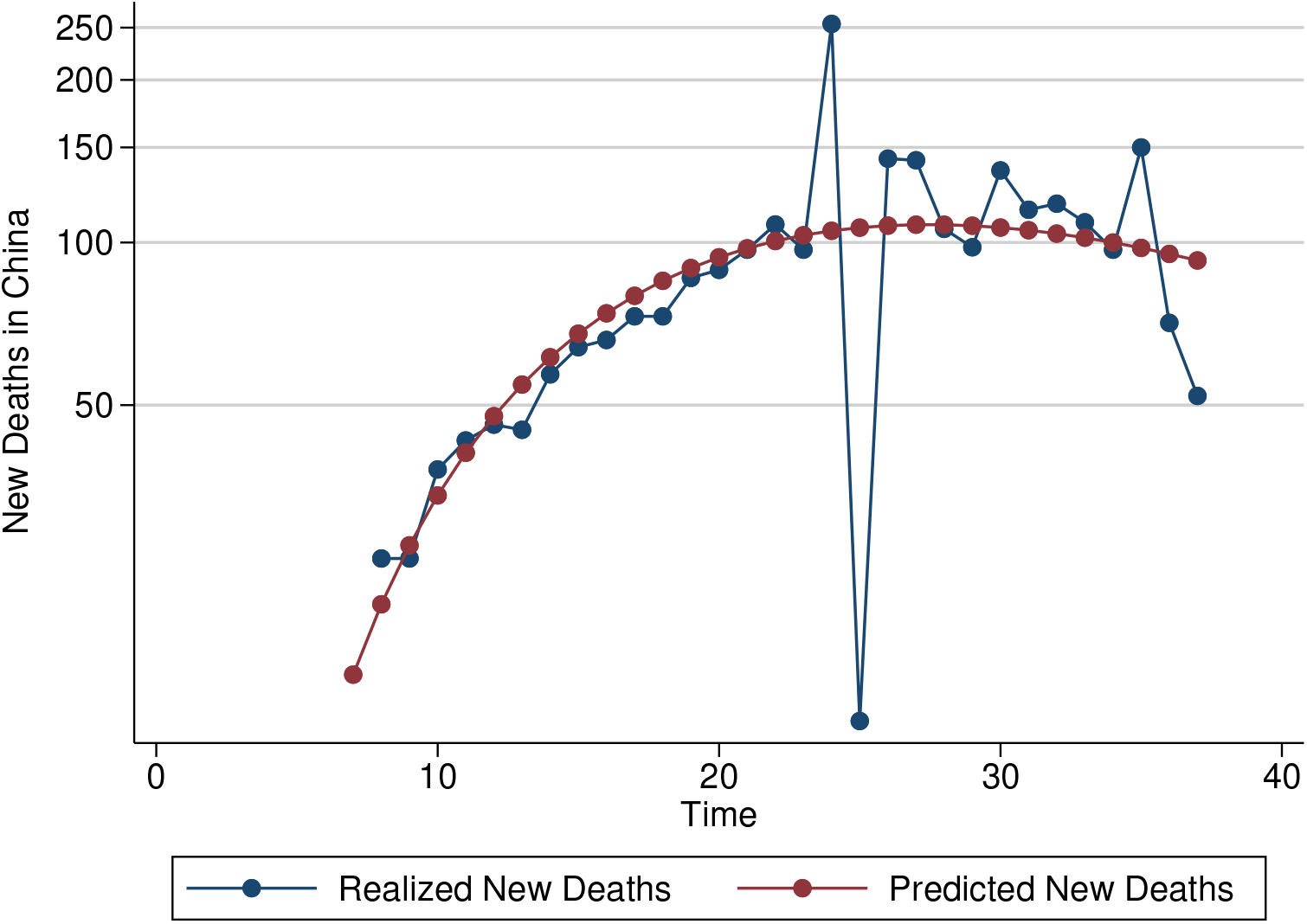
The daily number of deaths in China vs. Time (days since 1/21/2020) on a log-linear plot. The predicted new deaths is fit using the estimates reported in Tab. 2. Data from World Health Organization [2020].

The power-law (fractal) behavior that we postulated is related to the properties of the networks that carry out the propagation of the disease. Vazquez [2006] developed a network model in which the daily number of new cases *n*(*t*) in an epidemic follows a power-law with an exponential cutoff:

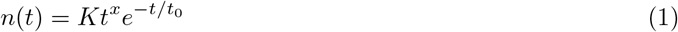

We have fit the data of China deaths to this function, and find *K* = 0.0854, *x* = 3.09, and *t*0 = 8.90 days (the time constant of decay), and this fit is shown in Fig. 2. Note here that the exponent (3.09) is somewhat above the value 2.28 that we were finding by a fit of a simple power law.

This can be integrated to find the total number of deaths as a function of time, in terms of the Gamma function Γ(*x*) and the incomplete Gamma function Γ(*a, x*):

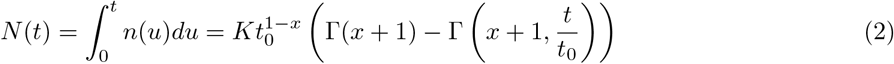

This plot is shown in Fig. 2, and we see a fairly good fit to the data.

The number of deaths outside of China has been growing rapidly. For several days it has been growing nearly exponentially, doubling every two to three days, but doe now appear to have slowed down a bit(although there are questions whether accurate data is being reported from one of the major hotspots, Iran). In Fig. 3, we show a log-linear plot of the deaths for the past 6 days, along with the deaths in China shifted by 31 days, and one can see somewhat similar behavior but fortunately with a downturn recently. We cannot predict any ultimate behavior for this behavior, but perhaps it will soon enter a power-law growth regime, before it enters the final stages.

**Figure 3:**
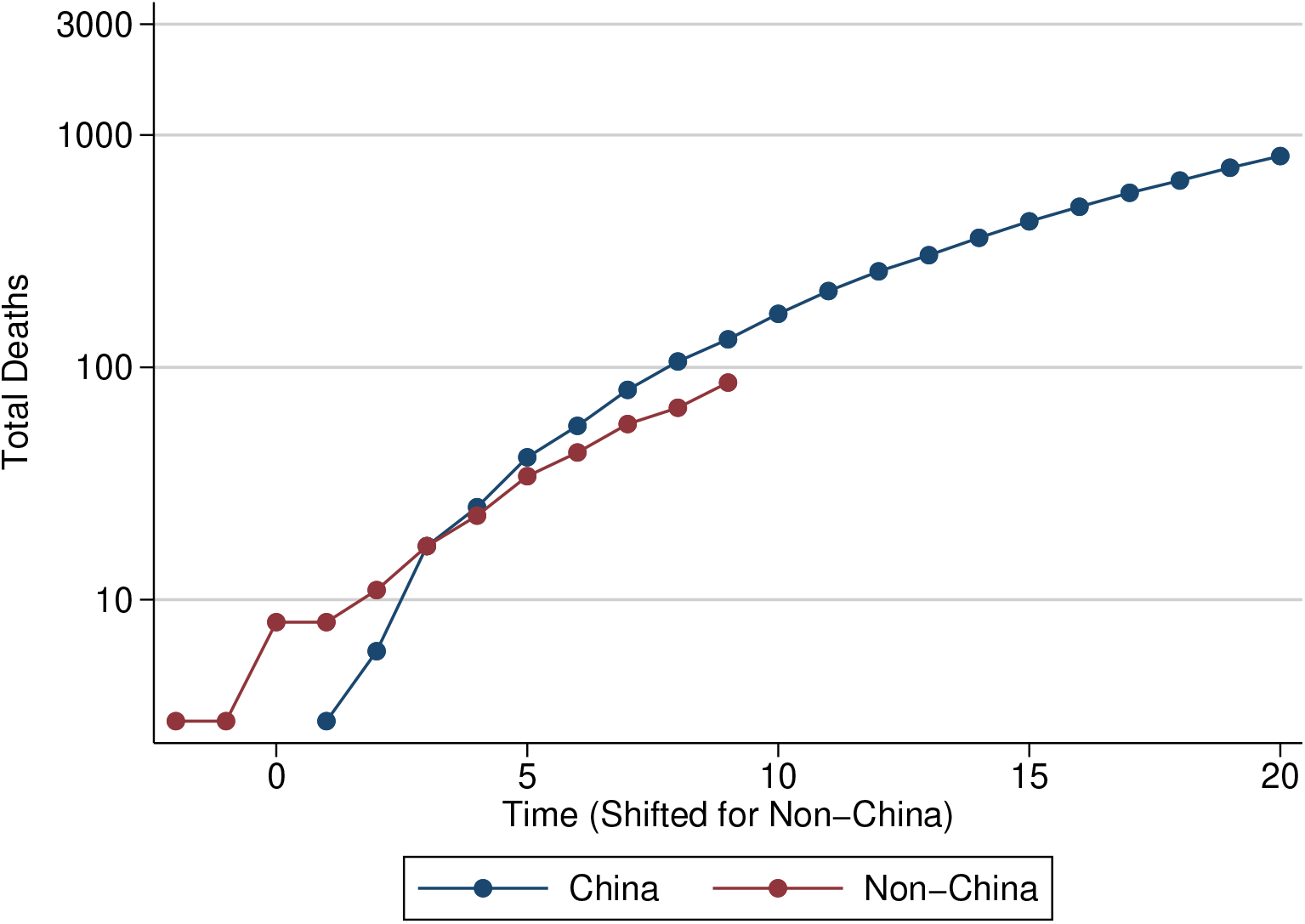
Total Deaths vs. Time, China and non-China on a log-linear plot. To align the trends, we subtract 31 from the timing of the data points from outside of China. Data from World Health Organization [2020]. Data from non-China cases up to March 1, 2020.

It is likely, with the normal international travel in the months of December and January, that the virus has been spread to many parts of the world, and cases have not been properly diagnosed. Thus we expect additional flair-ups to appear in different places in the world. But hopefully, with better understanding and appreciation of the disease, and appropriate responses, these flare-ups will be contained and a global pandemic can be averted.

Since we wrote our paper (and before as well), numerous papers have appeared modeling the epidemiology of the disease.

Li et al. [2020] confirm the power-law behavior we proposed for deaths in China, and also find power-law behavior (but with different exponents) for the number of infections and the number of recoveries. Fitting the number of cases to a quadratic on the log-log plot, they predicted an effective end of the epidemic around March 3 with a total of 83,000 cases, which so far has been well born out.

Numerous papers have considered compartment models and other methods to model the evolution of the epidemic Li [2020], Gamero et al. [2020], Rabajante [2020], Li and Feng [2020], Peng et al. [2020], Chen et al. [2020], Hu et al. [2020], Maier and Brockmann [2020], Li et al. [2020], Zhou et al. [2020], Xu et al. [2020], Liu et al. [2020]. These tend to predict an S-shaped curve with a tapering off in the near future as is being seen. These models depend upon assumptions of the reproduction rate, incubation period, etc.

The authors thank Greg Huber, Ginestra Bianconi, and Youjin Deng for discussions and communications related to this work.

## 2 Original article

The number of cases of novel coronavirus 2019 n-CoV (which has recently been renamed as COVID-19) that started in China less than three months ago has been growing rapidly, pointing to a high infectivity. The mortality rate seems to fall somewhere between the common flu (0.1%) and the SARs coronavirus (10%). In Figs. 4a and 4b, we plot the total number of confirmed cases and total number of reported deaths, as a function of the number of days beginning with January 21, 2020, when the WHO first started reporting on this pandemic in its Situation Reports [World Health Organization, 2020]. Both cases and deaths continue to grow at an alarming rate.

**Figure 4:**
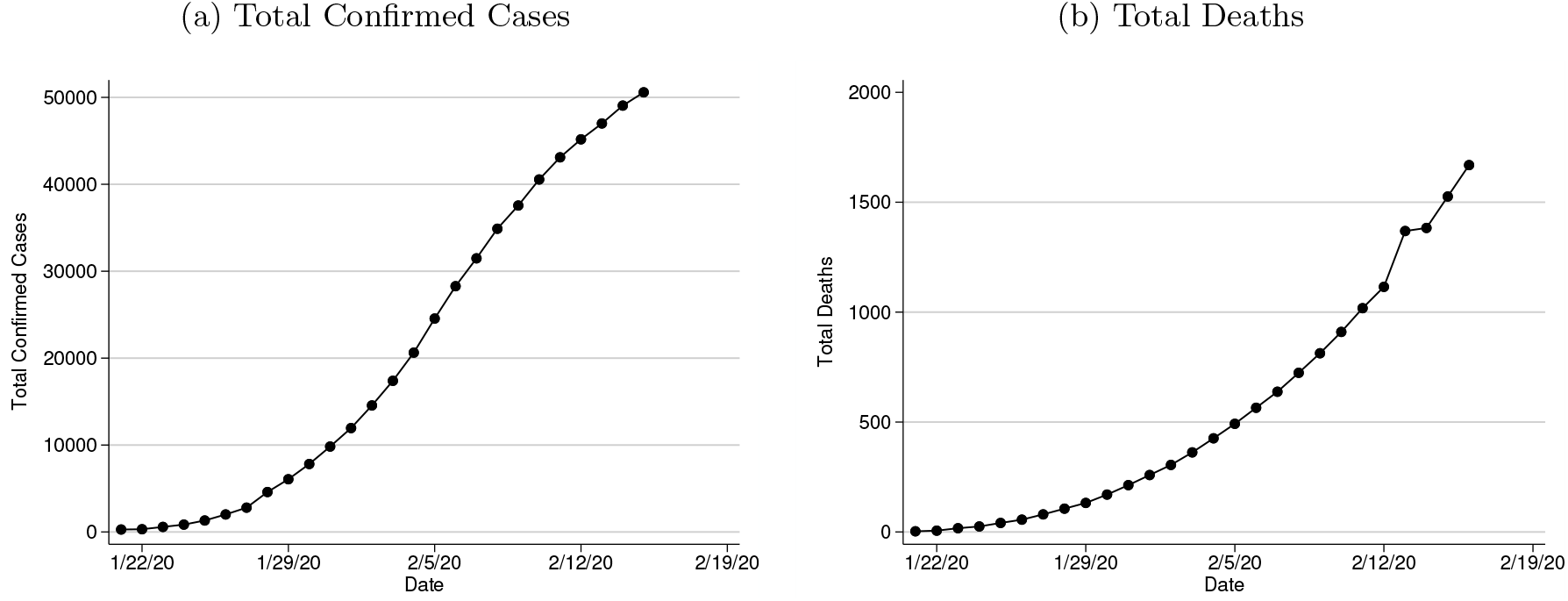
Raw Trends in COVID-19 Cases and Deaths. Note: Data from World Health Organization [2020].

In standard epidemiological analysis, one assumes that the number of cases in diseases like this one grows exponentially, based upon the idea of a fixed reproduction rate. If each person infects *n* other people where *n >* 1 (the reproduction number), then the total number of cases should grow as *nt/τ* = *e*^*at*^, where *τ* is the incubation time, which depends on the characteristics of the particular disease. Several studies of COVID-19 have taken this approach to model the number of cases [e.g., Zhao et al., 2020]. This assumes there is no inhibition due to the interaction with already infected people, quarantine, or other prophylactic measures.

While the data display large growth, they do not in fact follow exponential behavior. In the inset in Fig. 5 we show a log-normal plot of the mortality data, and the behavior is clearly not linear as it would be for exponential growth. In contrast, we find that the data are very well fit by assuming a power-law behavior with an exponent somewhat greater than two, as shown in the log-log plot in Fig. 5. The non-integer value of this power-law suggests a fractal type of behavior of the susceptibility mechanism, as we discuss below.

**Figure 5:**
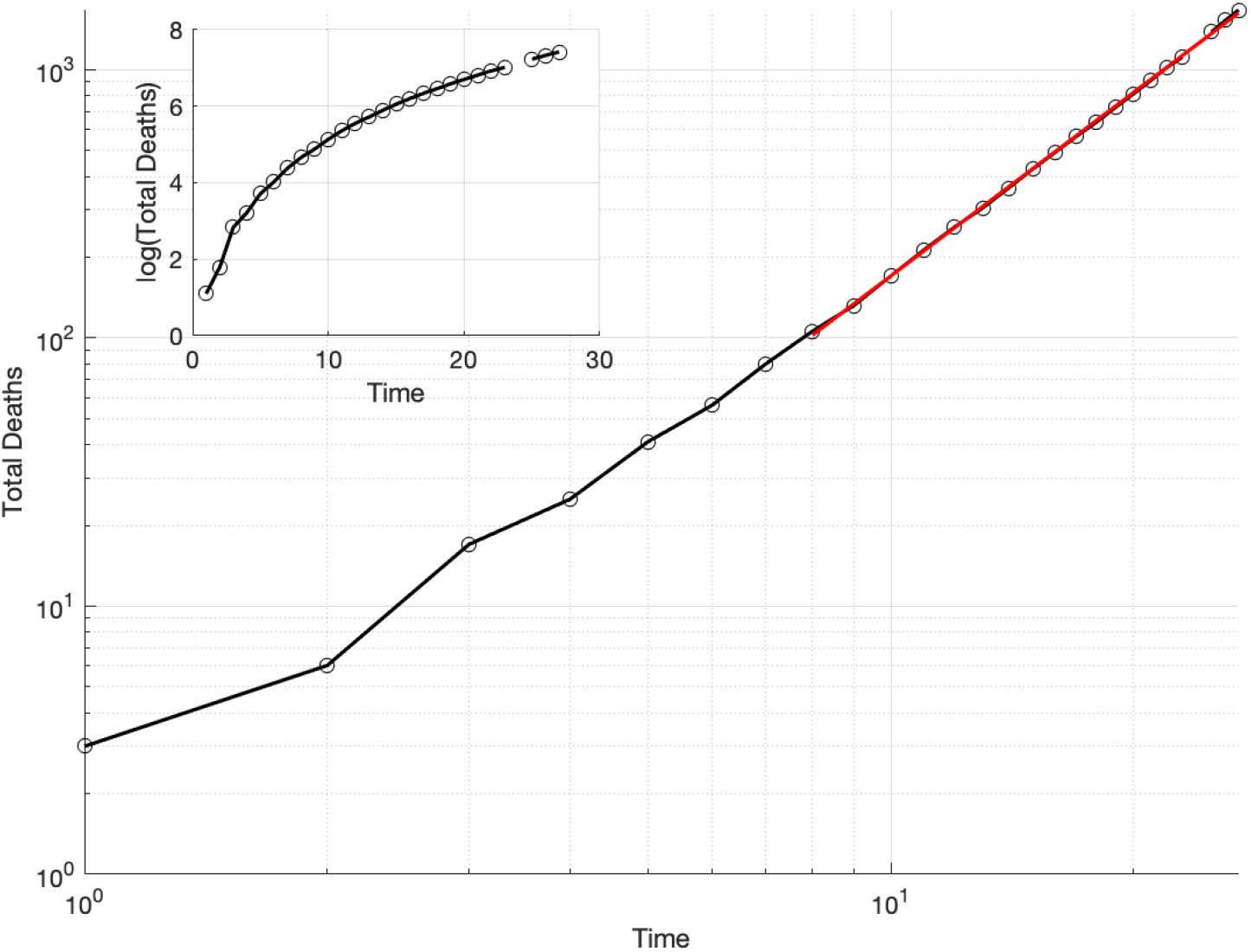
Total Deaths vs. Time (days since 1/21/2020) on a log-log plot (main plot) and log-normal plot (inset) Note: The red line in the large plot corresponds to the power-law fit for periods 8–27, omitting 24, i.e., 1/28/2020 to 2/15/2020, omitting spurious data for 2/13/2020. Data from World Health Organization [2020].

First we notice that the total number of deaths in Fig. 4b appear to be growing in a roughly quadratic form (see Tab. 1 in Appendix A for the estimates of the quadratic fit), while the number of confirmed cases shown in Fig. 4a appears to follow similar behavior but is slowing down in the last few data points. There could be measurement error due to multiple factors.^1^ Starting on February 7, those individuals who test positive for COVID-19 but show no symptoms are no longer being reported as confirmed cases [Cohen, 2020]. We focus our analysis on deaths to circumvent some of the ambiguities of reporting confirmed cases, assuming that deaths are a more reliable indication of the extent of the disease. (However, there are reports that some deaths might be reported as due to other causes, such as severe pneumonia, and not attributed to this virus, so these numbers may also be inaccurate. Still, one set of data should give a good indication of the general trend of the pandemic.)

Growing as *t*^2^ is what one would expect if a disease were growing in a population at the periphery of a compact region of infection. We believe that this is more likely than an exponential growth, because the number of susceptible individuals around an infected individual decays with time. Individuals already infected might face increased immunity and there are other individuals who might have had a mild infection that imparts immunity but without developing symptoms warranting testing. These various effects would inhibit the exponential growth of the virus.

We find that a power-law provides a better fit to the data than a simple quadratic, as postulated by Brandenburg [2020]. In Fig. 5 we plot the number of total deaths vs. time on a log-log plot. As Fig. 6 shows, considering all of the time periods (excluding report #24) from January 21 to February 15 (1–27 on the horizontal axis), the estimated coefficient is statistically indistinguishable from 2. However, removing just the first period period (2–27 on the horizontal axis), the estimated coefficient is statistically larger than 2. As more periods are removed, the coefficient moves farther away from 2. Even at this preliminary stage with few data points, this illustration suggests that a power-law fit is more appropriate than a quadratic one.

**Figure 6:**
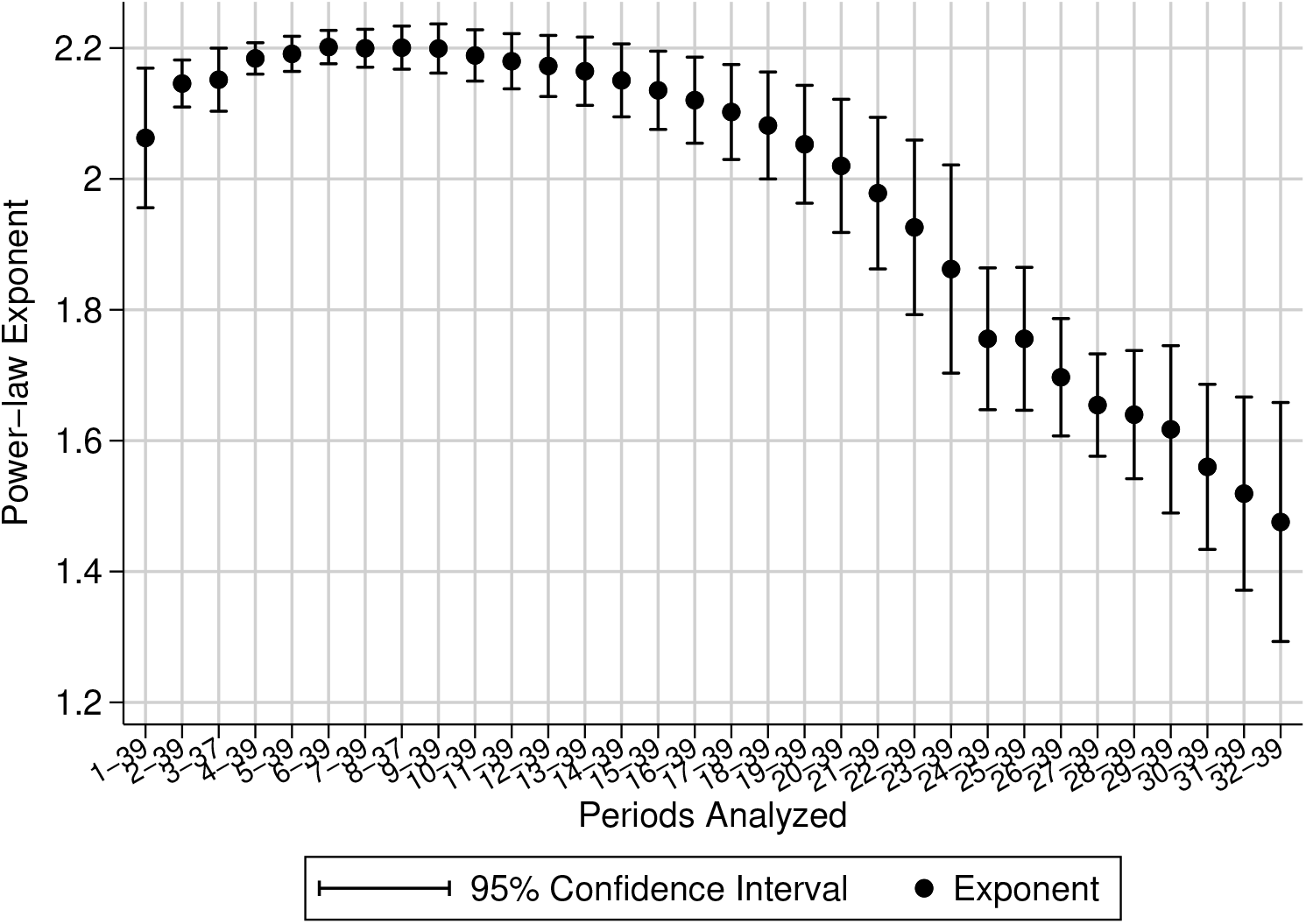
Power-law Coefficients, Dropping Earlier Periods. Note: The coefficients are the ordinary least squares estimates of *β*_1_ in the regression log(Deaths) = *β*_0_ + *β*_1_ log(time) + *ε* where *ε* is unobserved and assumed to be time-independent and distributed according to a *t* distribution with *n −* 2 degrees of freedom. Robust standard errors are calculated for suggestive inference, considering the small sample size. Data from World Health Organization [2020].

For illustration purposes, consider periods 8–27. The estimated coefficient is approximately 2.27 (the bootstrapped standard error with 5,000 replications is 0.014). This would imply that as the number of days of the pandemic increases by a factor of 2^1*/*2.27^ = 1.357, the number of cases will double. Thus, starting from day 27 (Feb. 16) with 1,669 deaths, the number of deaths will double to approximately 3,340 on day 37 (Feb. 22), and then double again to 6,680 on day 50 (March 10), and double again to 13,350 on day 67 (March 27). This is a large number of fatalities, but not nearly as large as if the growth were exponential. On the other hand, it is larger than some epidemiological models have been predicting.

The appearance of a fractional exponent and power-law behavior suggests an underlying fractal process. Based upon a kind of small-world interaction network where individuals have many local neighbors and occasional long-range connections (such as caused by people traveling on trains, boats and planes). The results imply that this network has an effective minimum path fractal dimension of 2.25. The implication is that the disease produces flares in which the growth is momentarily exponential, but then slows down until another flare-up. The averaged effect of this growth apparently yields a power-law.

Wu et al. [2020] suggest that if the pandemic spreads into new cities, exponential growth like what happened in Wuhan in the early stages would occur. Hopefully, if this were the case, it would be quickly stopped and slower behavior would ensue. But our prediction of approximately 13,350 deaths by March 27 relies on the assumption that a new major outbreak will not happen. Hermanowicz [2020] and Roosa et al. [2020] have recently modeled the disease using a logistical and other growth models. These models are able to predict the ultimate number of cases, and find results in the range of 20,000 to 60,000. We believe the numbers will be much greater, and expect a number of *deaths* in this range, if the fractal growth continues as it has for a few more months.

We have followed our prediction for several days and have found no marked change in behavior, except for the data from WHO Situation Report #24, which gave an outsized jump of 254 cases to a total of 1,369 deaths. Reports stated that the data from China were incorrect that day because of double-counting of the results, but so far the WHO has not revised report #24. Looking at the large deviation from our prediction in fact led us to suspect that that data point was an outlier at the time.

In recent days there has been a drop-off in the growth of new confirmed cases from China, suggesting an attenuation of the pandemic. We do not see that drop-off in the death rate. There are two possible explanations for this discrepancy: (1) the number of confirmed cases may be showing the variability as it has in the past, and the current numbers are not capturing the actual situation, or (2) the pandemic is indeed slowing down but it will take a while (perhaps a week) for that to show up in the fatality data. Of course, we hope the latter is the case, but are not entirely confident considering the history of the reporting problems concerning this pandemic.

Deviations above this power-law behavior might indicate that the pandemic is expanding from the current level of control, while deviations below might indicate that the disease is starting to fade. This analysis is early in the outbreak of COVID-19 and cannot predict the length of the outbreak nor the final fatality. However, it suggests that under current conditions, power-law growth better approximates the number of deaths than exponential growth, and may provide a tool to monitor whether a fundamental change is occurring. Hopefully, continued vigilance, timely testing, and careful care will end this scourge on humanity.

The authors report no funding related to this research and have no conflicting financial interests.

## Data Availability

All data comes from the World Health Organization site, and all statistical calculation data are included as Supplementary Files

## A Statistical Testing

Table 1 lists the estimated coefficients from the regression Deaths = *β*_0_ + *β*_1_time + *β*_2_*time*^2^ + *ε*, where *ε* is assumed to be time-independent and Normally distributed. We also report bootstrapped standard errors in brackets, which are more conservative in this small-sample context. See Efron and Tibshirani [1994] and Horowitz [2001] for introductions to the bootstrap method.

**Table 1:** Estimates of a Quadratic Model of Deaths

|  |  |
| --- | --- |
| Time | 1.954<br>(4.065)<br>[4.504] |
| Time<sup>2</sup> | 2.123***<br>(0.122)<br>[0.132] |
| Constant | -39.46<br>(23.84)<br>[29.23] |
| Observations | 36 |
| $R^2$ | 0.996 |

Note: Parametric standard errors are in parentheses, bootstrapped standard errors are in square brackets. We perform 5,000 replications. Data from World Health Organization [2020]. \* ( $p < 0.05$ ), \*\* ( $p < 0.01$ ), \*\*\* ( $p < 0.001$ ).

In Fig. 6, the confidence intervals are calculated as follows. First, robust standard errors are calculated to account for heteroskedasticity. These standard errors are more conservative, which is advantageous considering our small sample size. Next, we calculate the critical value (the 95th quantile) from a *t*-distribution with *n* 2 degrees of freedom. The value *n* changes with each point along the *x*-axis. As fewer periods are considered, the critical value increases. The degrees of freedom is a function of this *n*, but subtracting two to account for the fact that two parameters are estimated (*β*_0_, *β*_1_). The standard errors are quite small for these calculations. Considering periods 2-27 produces a bootstrapped standard error of 0.033, using 5,000 bootstrap replications.

Fig. 7 shows the estimated power-law exponents starting at period 8 and adding more periods forward. This mirrors our process of testing this model as more data were published on the number of deaths. The coefficient is stable. The confidence intervals are calculated analogously to those in Fig. 6. Note that data at period 24 are omitted due to probable inaccuracy, as we discuss above.

**Figure 7:**
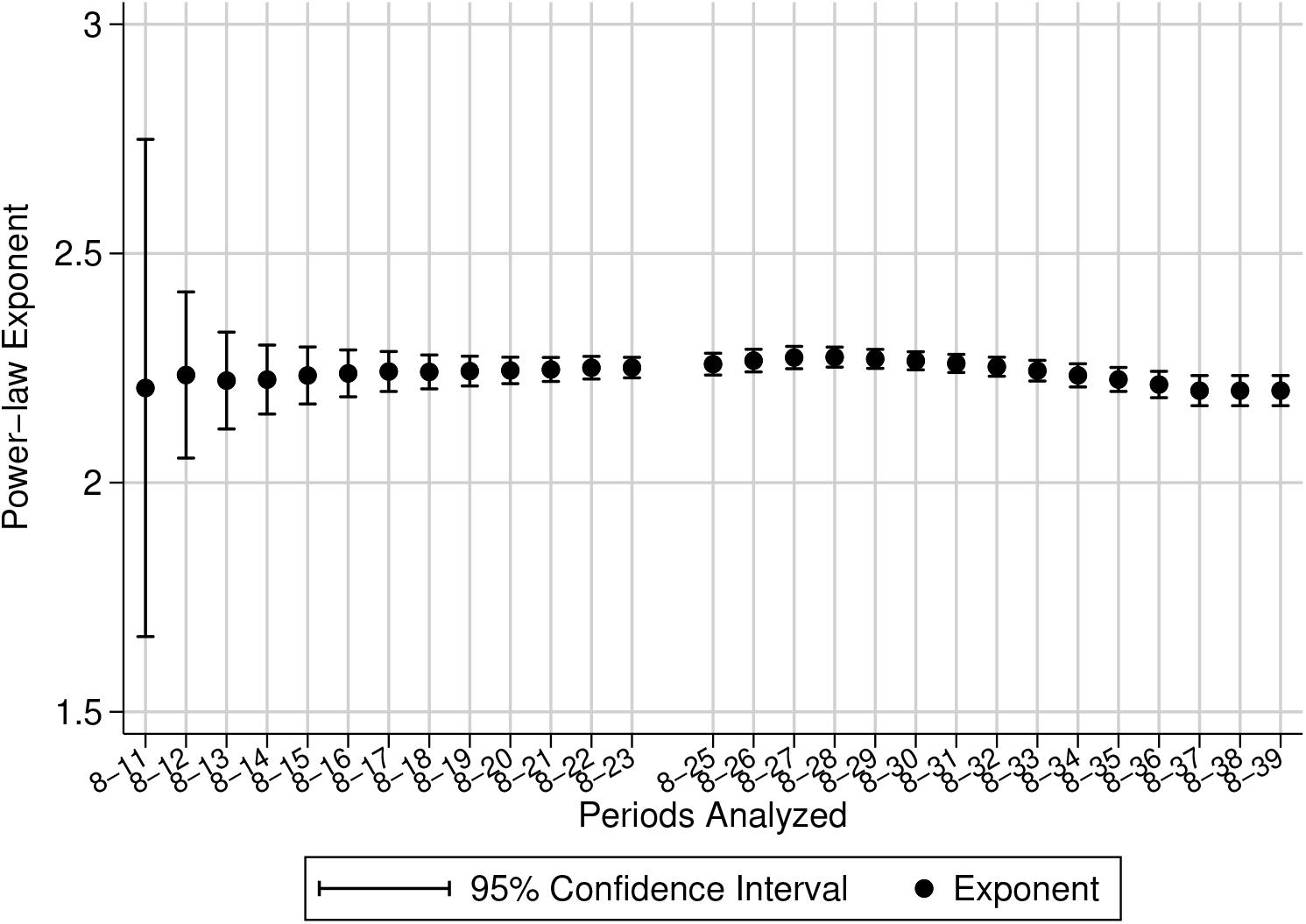
Power-law Coefficients, Dropping Later Periods. Note: The coefficients are the ordinary least squares estimates of *β*_1_ in the regression log(Deaths) = *β*_0_ + *β*_1_ log(time) + *ε* where *ε* is unobserved and assumed to be time-independent and distributed according to a *t* distribution with *n −* 2 degrees of freedom. Robust standard errors are calculated for suggestive inference, considering the small sample size. Data from World Health Organization [2020].

Tab. 2 displays the estimates of the mixed power law and exponential model. Given that the disease has been in China the longest, we estimate perform the estimation on only China. We exclude periods 24 and 25 (2/13/2020 and 2/14/2020) to estimate the coefficients. The estimates are significant.

**Table 2:** Estimates of a Mixed Power Law and Exponential Model

|  |  |
| --- | --- |
| $x$ | 3.089***<br>(0.441)<br>[0.576] |
| $-1/t_0$ | -0.113***<br>(0.0221)<br>[0.0308] |
| $\log(K)$ | -2.460**<br>(0.850)<br>[1.087] |
| Observations | 28 |
| $R^2$ | 0.844 |

Note: This table displays the estimates from Eq. 1. Parametric standard errors are in parentheses, bootstrapped standard errors are in square brackets. We perform 5,000 replications. Data from World Health Organization [2020].
\*( $p < 0.05$ ), \*\* ( $p < 0.01$ ), \*\*\* ( $p < 0.001$ ).

The replication files include a `.xlsx` file of the compiled reports from World Health Organization [2020]. These data are publicly available, but we condense the information used in this report. A `.do` file contains the STATA code to clean the data, perform the statistical analysis, and produce the plots. A machine-specific environment variable needs to be set with the accompanying file structure. Once this is set, the code will run seamlessly on any machine with STATA installed. The Matlab file is only used to create a graph.

## Footnotes

1 Some of these factors includes misrepresentation of the number of confirmed cases, and varying the criteria used for the identification of the disease. These have been discussed in news reporting [Wang, 2020, BBC, 2020]. This analysis does not aim to quantify any intentional bias in the reported data.

## References

BBC. Coronavirus: “Way too early” to predict end of outbreak, WHO says, February 2020. URL https://www.bbc.com/news/world-asia-china-51482994.

Axel Brandenburg. Quadratic growth during the 2019 novel coronavirus epidemic. 2002.03638 [q-bio], February 2020. URL http://arxiv.org/abs/2002.03638. 2002.03638.

Baoquan Chen, Mingyi Shi, Xingyu Ni, Liangwang Ruan, Hongda Jiang, Heyuan Yao, Mengdi Wang, Zhenghua Song, Qiang Zhou, and Tong Ge. Data Visualization Analysis and Simulation Prediction for COVID-19. 2002.07096 [physics, q-bio], February 2020. URL http://arxiv.org/abs/2002.07096. 2002.07096.

Jon Cohen. Scientists are racing to model the next moves of a coronavirus that’s still hard to predict, February 2020. URL https://www.sciencemag.org/news/2020/02/scientists-are-racing-model-next-moves-coronavirus-thats-still-hard-predict.

Bradley Efron and Robert J. Tibshirani. An Introduction to the Bootstrap. CRC Press, 1994.

Javier Gamero, Juan A. Tamayo, and Juan A. Martinez-Roman. Forecast of the evolution of the contagious disease caused by novel coronavirus (2019-nCoV) in China. 2002.04739 [q-bio, stat], February 2020. URL http://arxiv.org/abs/2002.04739. 2002.04739.

Slav W Hermanowicz. Forecasting the Wuhan coronavirus (2019-nCoV) epidemics using a simple (simplistic) model - update (Feb. 8, 2020). Epidemiology, In Press, February 2020. doi :10.1101/2020.02.04.20020461. URL http://medrxiv.org/lookup/doi/10.1101/2020.02.04.20020461.

Joel L. Horowitz. The Bootstrap. In Handbook of Econometrics, volume 5, pages 3159–3228. Elsevier, 2001. ISBN 978-0-444-82340-3. doi :10.1016/S1573-4412(01)05005-X. URL https://linkinghub.elsevier.com/retrieve/pii/S157344120105005X.

Zixin Hu, Qiyang Ge, Li Jin, and Momiao Xiong. Artificial Intelligence Forecasting of Covid-19 in China. 2002.07112 [q-bio], February 2020. URL http://arxiv.org/abs/2002.07112. 2002.07112.

Jun Li. A Robust Stochastic Method of Estimating the Transmission Potential of 2019-nCoV. 2002.03828 [physics, q-bio, stat], February 2020. URL http://arxiv.org/abs/2002.03828. 2002.03828.

Ming Li, Jie Chen, and Youjin Deng. Scaling features in the spreading of COVID-19. 2002.09199 [physics, q-bio], February 2020. URL http://arxiv.org/abs/2002.09199. 2002.09199.

Qiang Li and Wei Feng. Trend and forecasting of the COVID-19 outbreak in China. 2002.05866 [q-bio], February 2020. URL http://arxiv.org/abs/2002.05866. 2002.05866.

Zhihua Liu, Pierre Magal, Ousmane Seydi, and Glenn Webb. Predicting the cumulative number of cases for the COVID-19 epidemic in China from early data. 2002.12298 [math, q-bio], February 2020. URL http://arxiv.org/abs/2002.12298. 2002.12298.

Benjamin F. Maier and Dirk Brockmann. Effective containment explains sub-exponential growth in confirmed cases of recent COVID-19 outbreak in Mainland China. 2002.07572 [physics, q-bio], February 2020. URL http://arxiv.org/abs/2002.07572. 2002.07572.

Liangrong Peng, Wuyue Yang, Dongyan Zhang, Changjing Zhuge, and Liu Hong. Epidemic analysis of COVID-19 in China by dynamical modeling. 2002.06563 [q-bio], February 2020. URL http://arxiv.org/abs/2002.06563. 2002.06563.

Jomar F. Rabajante. Insights from early mathematical models of 2019-nCoV acute respiratory disease (COVID-19) dynamics. 2002.05296 [q-bio], February 2020. URL http://arxiv.org/abs/2002.05296. 2002.05296.

K. Roosa, Y. Lee, R. Luo, A. Kirpich, R. Rothenberg, J.M. Hyman, P. Yan, and G. Chowell. Real-time forecasts of the 2019-nCoV epidemic in China from February 5th to February 24th, 2020. Infectious Disease Modelling, February 2020. ISSN 24680427. doi :10.1016/j.idm.2020.02.002. URL https://linkinghub.elsevier.com/retrieve/pii/S2468042720300051.

Alexei Vazquez. Polynomial growth in age-dependent branching processes with diverging reproductive number. Physical Review Letters, 96(3):038702, January 2006. ISSN 0031-9007, 1079-7114. doi :10.1103/PhysRevLett.96.038702. URL http://arxiv.org/abs/cond-mat/0505116arxivcond-mat/0505116.

Vivan Wang. How Many Coronavirus Cases in China? Officials Tweak the Answer, February 2020. URL https://www.nytimes.com/2020/02/12/world/asia/china-coronavirus-cases.html.

World Health Organization. Novel Coronavirus (2019-nCoV) situation reports. Technical Report 1-24, WHO, January 2020. URL https://www.who.int/emergencies/diseases/novel-coronavirus-2019/situation-reports.

Joseph T. Wu, Kathy Leung, and Gabriel M Leung. Nowcasting and forecasting the potential domestic and international spread of the 2019-nCoV outbreak originating in Wuhan, China: a modelling study. The Lancet, In Press, Jan 2020. ISSN 01406736. doi :10.1016/S0140-6736(20)30260-9. URL https://linkinghub.elsevier.com/retrieve/pii/S0140673620302609.

Xiaowei Xu, Xiangao Jiang, Chunlian Ma, Peng Du, Xukun Li, Shuangzhi Lv, Liang Yu, Yanfei Chen, Junwei Su, Guanjing Lang, Yongtao Li, Hong Zhao, Kaijin Xu, Lingxiang Ruan, and Wei Wu. Deep Learning System to Screen Coronavirus Disease 2019 Pneumonia. 2002.09334 [physics], February 2020. URL http://arxiv.org/abs/2002.09334. 2002.09334.

Shi Zhao, Qianyin Lin, Jinjun Ran, Salihu S Musa, Guangpu Yang, Weiming Wang, Yijun Lou, Daozhou Gao, Lin Yang, Daihai He, and Maggie H Wang. Preliminary estimation of the basic reproduction number of novel coronavirus (2019-nCoV) in China, from 2019 to 2020: A data-driven analysis in the early phase of the outbreak. International Journal of Infectious Diseases, In Press, January 2020. ISSN 1201-9712. doi :10.1016/j.ijid.2020.01.050. URL http://www.sciencedirect.com/science/article/pii/S1201971220300539.

Yimin Zhou, Zuguo Chen, Xiangdong Wu, Zengwu Tian, Liang Cheng, and Lingjian Ye. The Outbreak Evaluation of COVID-19 in Wuhan District of China. 2002.09640 [physics, q-bio], February 2020. URL http://arxiv.org/abs/2002.09640. 2002.09640.

